# Indication for SARS-CoV-2 serology: first month follow-up

**DOI:** 10.1101/2020.06.30.20140715

**Authors:** Alix T. Coste, Katia Jaton, Matthaios Papadimitriou-Olivgeris, Antony Croxatto, Gilbert Greub

**Author notes:** **Corresponding author: Professeur Gilbert GREUB** - MD-PhD. Institut de microbiologie de l’Université de Lausanne Bugnon 48, CH-1011 Lausanne, Switzerland. Participated equally to the study (Antony Croxatto, Gilbert Greub).

## Abstract

SARS-CoV-2 detection is mainly performed by RT-PCR but recently serological tests were made available. A first one month follow-up of the SARS-CoV-2 serology records was performed in our laboratory to precise the diversity and proportion of the SARS-CoV-2 serology test indications and to identify new valid indications (meningoencephalitis, vasculitis, …)

## Introduction

A new coronavirus was discovered late in 2019 following an unusual number of pneumonia in the city of Wuhan in China. This virus was named SARS-CoV-2 and the disease was named Coronavirus Disease 2019 (CoVID-19) (https://www.who.int/news-room/detail/27-04-2020-who-timeline---covid-19). This pandemic arrived in Switzerland on 25^th^ February 2020. On 18^th^ May 2020, as many as 31’218 persons were documented with a SARS-CoV-2 infection causing 1956 deaths in Switzerland, whereas in Vaud county, where our university hospital is located, 5413 cases were documented with 420 deaths (www.corona-data.ch). Detection of the SARS-CoV-2 was mainly performed by PCR on nasopharyngeal swab during the acute phase of the disease [1, 2]. Recently, commercial serological tests were available allowing the determination of the serostatus of patients [3]. The SARS-CoV-2 serology was implemented and used for patients care since 14^th^ April 2020 in our laboratory of microbiology located at the “Centre Hospitalier Universitaire Vaudois” (CHUV).

Initially, only few indications were recognized by public health authorities, but we rapidly extended these indications to include the list detailed in Table 1. Our laboratory performed a prospective surveillance of the SARS-CoV-2 serologic test requested during the first 5 weeks, by specifically looking at rate of the different accepted indications. We also prospectively checked for a sudden increase in testing requests from a specific ward of our university hospital and/or from a tertiary hospital, a private clinic or a specific private practitioner. Finally, we also identified prospectively new valid indications.

**Table 1:**
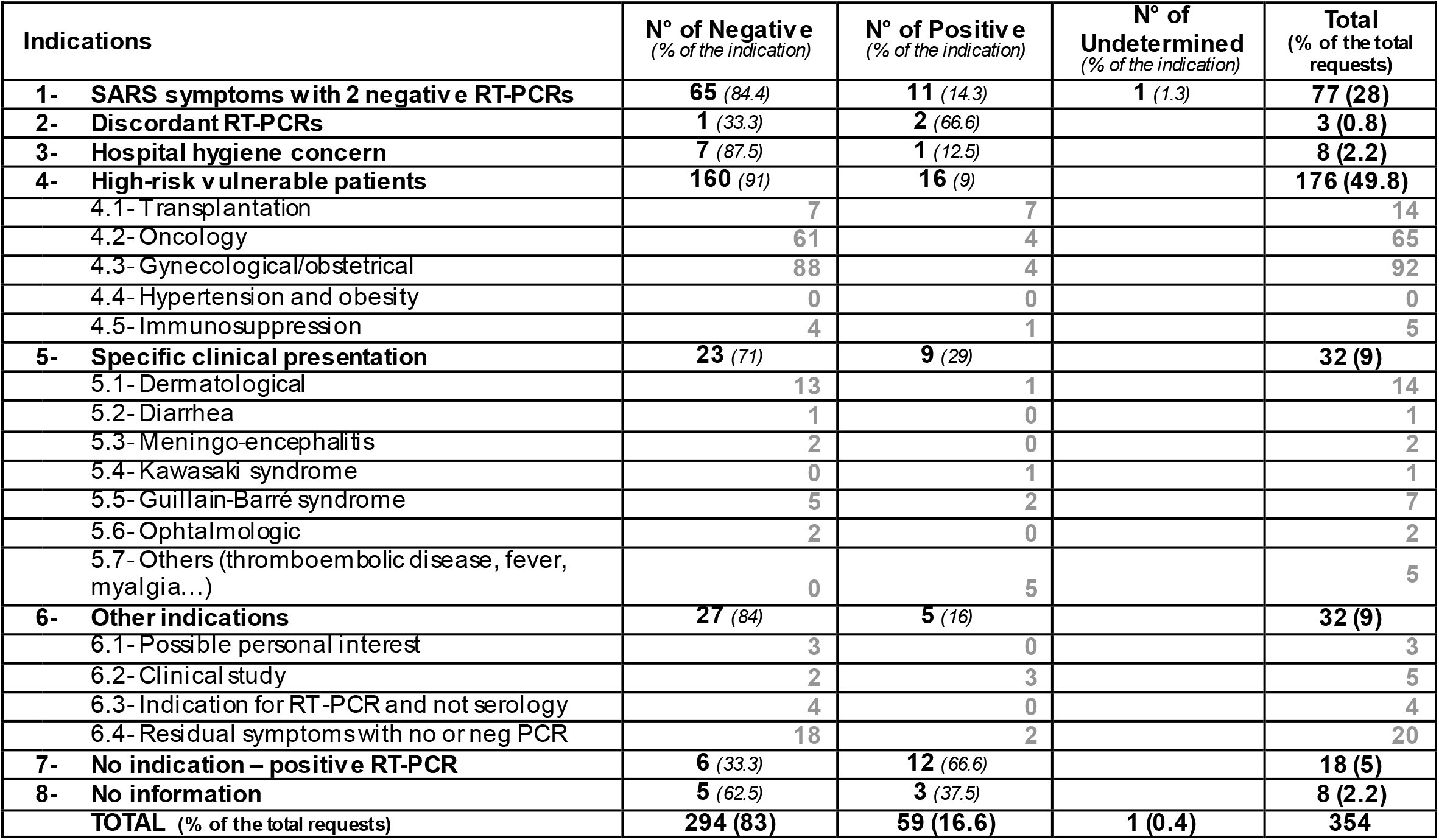
Clinical indications of the CHUV SARS-CoV-2 serology requests

The aims of this work were (i) to precise the diversity and proportion of indications for the SARS-CoV-2 serological test and (ii) to report any deviation of its usage.

## Results

From the 14^th^ April 2020 to 18^th^ May 2020, 686 SARS-CoV-2 serological tests were performed in our laboratory using the Epitope Diagnostics IgG ELISA (San Diego, CA). Twenty-nine (4%) were prescribed by private practitioners, 303 (44%) by hospital-based physicians working in tertiary hospitals or clinics, and 354 (52%) by physicians of our institution. These tests were performed on 602 patients, including 367 female subjects (61%) from 0 to 89 years old, and 235 male persons (39%) from 1 to 96 years old. On the 686 serological tests performed 159 were positive (23%), 522 were negative (76%), and five were undetermined (3%). Among the 28 serologies prescribed by private practitioners, five were positive (18%) and 23 were negative (82%). For the 303 serologies requested by hospital-based physicians working in tertiary hospitals or clinics, 94 were positive (31%), 205 were negative (68%), and four were undetermined (13%). Finally, for the 354 serological tests from clinicians working at CHUV, 59 were positive (17%), 294 were negative (83%) and one was undetermined (<1%). Initially, we observed that tertiary hospital B sent more serology analyses than expected. They had specific directives in their hospital to systematically perform a SARS-CoV-2 RT-PCR screening and serology to any patient arriving at the hospital. This practice being not in accordance with the county directives, we notified this to the hospital B physicians. Then, starting from week three, we did not observe any specific prescription-related pattern (Figure 1).

**Figure 1:**
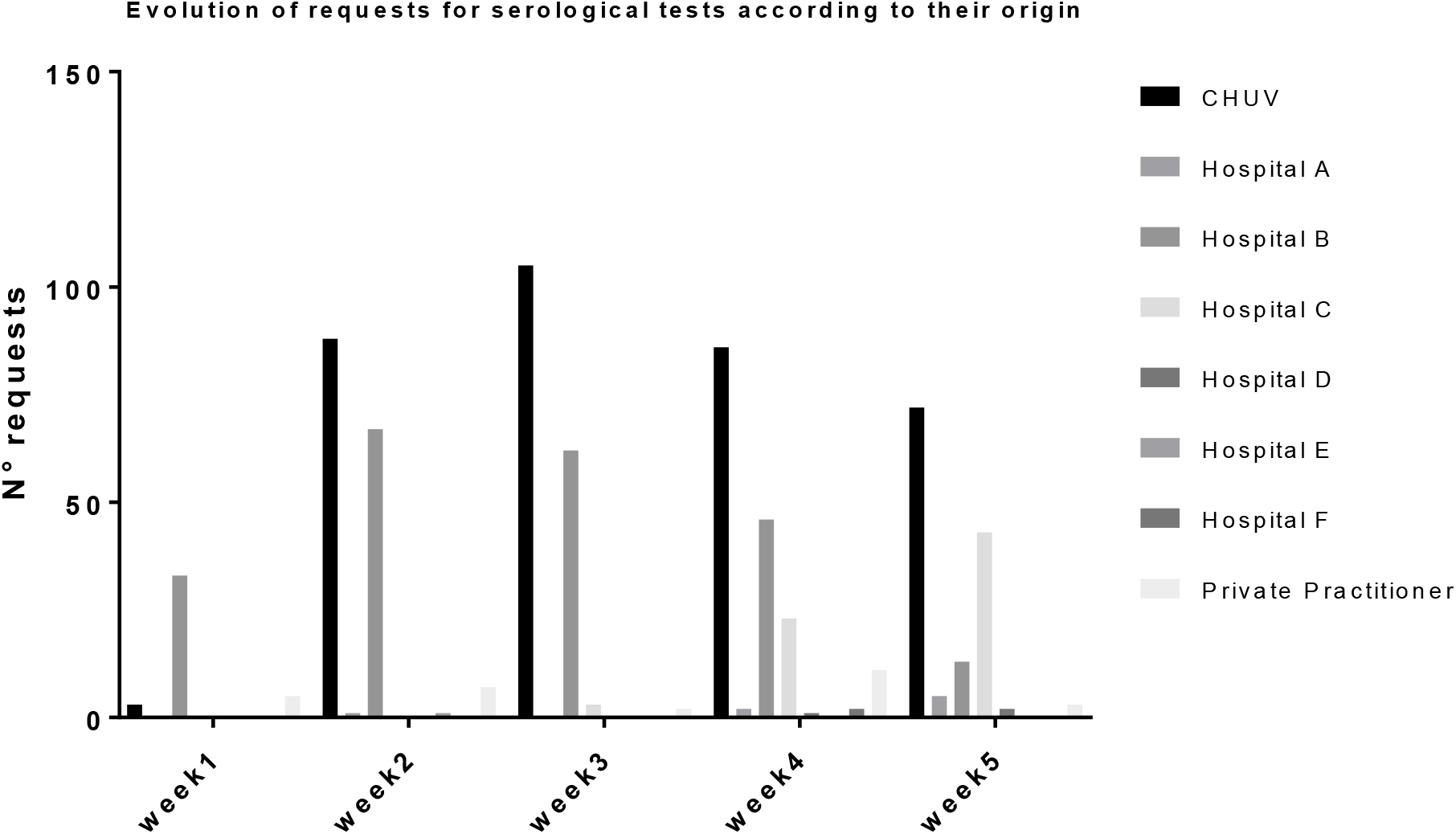
Evolution of the SARS-CoV-2 serology requests according medical center origin

Regarding the non-CHUV serological requests, we decided to focus only on the clinical information available on the laboratory request form. Only 22 (6.6%) of 332 serological requests contained such information. Fourteen serological tests were performed for patients with indication N°1 (SARS symptoms with ≥ 2 negative RT-PCR), one with indication N°2 (discordant RT-PCR), one with indication N°4.4 (vulnerable subjects: hypertension & obesity), three with indication N°5 (specific clinical presentation) including one with Guilla in-Barré syndrome, one with indication N°6.1 (personal interest) and two with indication N° 6.4 (residual symptoms but who had no RT-PCR results available). Noteworthy, there was no indication of serology for 22 (6.6%) of the 332 requested serologies, as they already had a positive SARS-CoV-2 RT-PCR, all coming from tertiary hospital B. This was due to the hospital B directive to systematically screen any incoming patient by RT-PCR and serology, to ideally increase the overall screening sensitivity. Noteworthy, the serology result can only be used to confirm an infection and not to assess the protection of the patient, since no clear relationship between the serostatus and the protection against a new infection by the SARS-CoV-2 was established. Therefore the serology was useless for these 22 cases, since a positive RT-PCR is clearly sufficient to prove the infection.

The 354 CHUV requests correspond to 313 patients. Among those 313 patients, 54 (17.2%) had a positive SARS-CoV-2 serology. A medical doctor recorded a date of symptoms onset for 42 of them. We could thus determine that the time between symptoms onset and serology requests was between 4 to 61 days, with a mean at 27 days, and a median at 25 days. On the 313 CHUV patients, 241 (77%) had a SARS-CoV-2 RT-PCR performed, including 46 (19%) with a positive RT-PCRs. On average, the serology was requested 10.8 days after the RT-PCR. This period is longer for 43 patients with a positive serology (20.8 days on average), and shorter for the 198 patients with a negative serology (8.9 days on average), as expected. Based on these results, when initially negative, we now systematically recommend the repetition of the serology two weeks later, especially when the first serology is done <15 days after symptoms onset. Indeed, in our hands, the Epitope Diagnostics test reached 96% sensitivity on sera taken ≥15 days after symptoms onset from hospitalize d patients with mild to severe symptoms [4].

For the 354 CHUV requests, an indication was determined from the electronic medical record of all patients except eight, and these indications, as well as their relative proportions and results are summarized in Figure 2. Clearly, the majority of the requests (49%) concerned vulnerable patients (indication N°4), regardless of their symptoms (Figure 2A). Among vulnerable patients, pregnant women represented about half of the requests (52%), followed by oncological cases (37%). Transplanted or other immunosuppressed patients represented only 8% and 3%, respectively, of the vulnerable subjects (Figure 2B). The second most frequent indication was SARS symptoms with two consecutive negative SARS-CoV-2 RT-PCRs (indication N°1), corresponding to 21% of the requests (Figure 2A). In third position are requests with either “specific clinical presentation” (indication N°5) or “other type of indications” (indication N°6; Figure 2). Among requests due to “specific clinical presentation”, patients of dermatology with skin vasculitis or endothelitis, and patients with suspected Guillain-Barré syndrome were the most frequent and represented 44% and 22% of these requests, respectively (Figure 2C). Interestingly, among requests for “other type of indications” (N°6), patients with residuals symptoms but without RT-PCR or a negative RT-PCR performed at the acute phase of the disease (indication N°6.4) represented 62.5% of this category, and 5% of all 354 requests (Figure 2D). The residuals symptoms with no documented or negative RT-PCR (indication N°6.4) appeared to be very important for patient care but was totally unexpected, since on 14^th^ April, when we started the SARS-CoV-2 serology, the occurrence of such post-infectious complications were not yet reported [5]. Today, such late complications appear to be common and typical of COVID-19 [6-8].

**Figure 2:**
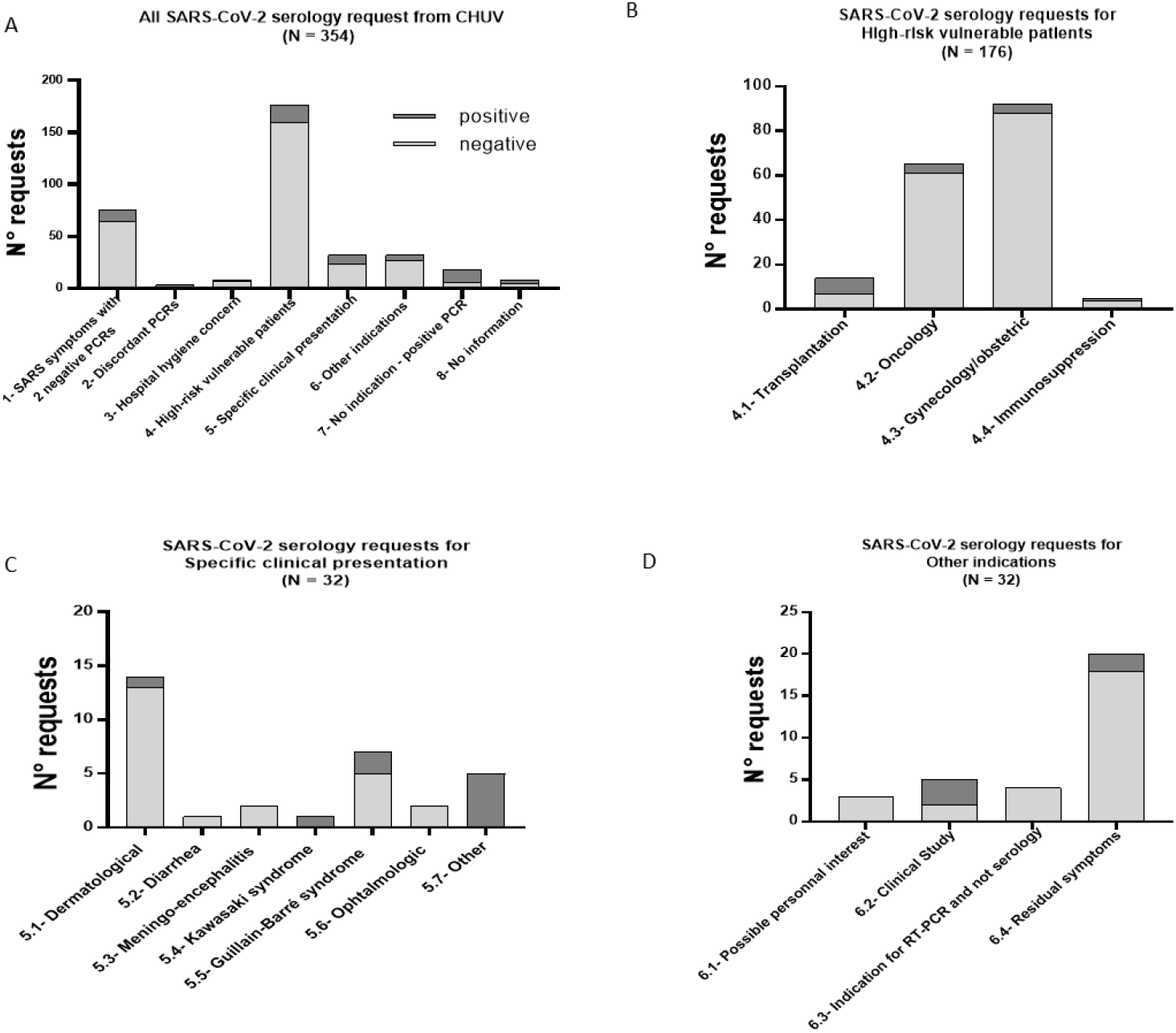
Repartition of the CHUV SARS-CoV-2 serology requests according clinical indication

One of the main limitation of this work is the short studied period, which do not include possible much later onset complications, necessitating a serological investigation. Some of the indications that we expected such as (i) investigation of thrombo-embolic events, and (ii) investigations of chronic fatigue syndrome were not recorded, likely due to the relatively small number of subjects, the lack of information for most serologies requested by private practitioners and the early timing of this work, as compared to the outbreak reducing the number of cases with late complications. Thus, this work should be extended over the next 6 to 12 months.

Overall, this work demonstrates the good adequacy of serological requests. Indeed, at least the in-hospita l doctors largely complied with in-house guidelines and did not sent serology requests only for scientific interest or personal interest. This work may serve as a seed for international guidelines regarding the indications of SARS-CoV-2 serology for patients care.

## Data Availability

All data are available on demand.

